# Cost Analysis of Vasopressin Use in Septic Shock and Predictive Performance of GOSSIS in ICU Mortality

**DOI:** 10.64898/2026.11.03.26367179

**Authors:** Kangho Suh, Shiyi Wang, Chenell L Donadee, Aimee Boeltz, Ian Barbash, Xiaojiang Guo, Saif Alam, Christopher M Horvat, Victoria Collier, Ryan M Rivosecchi

## Abstract

**Background:** Vasopressin is a commonly used adjunctive therapy in the management of septic shock. While vasopressin may provide clinical benefits, at times its acquisition cost has risen to being substantially more expensive than other second-line vasopressors such as epinephrine, phenylephrine, or dopamine. Additionally, mortality prediction tools such as the Global Open-Source Severity of Illness Score (GOSSIS) have emerged as a valuable resource to guide resource allocation in critical care. This study aimed to evaluate the healthcare costs associated with vasopressin use and assess the predictive performance of GOSSIS in patients with septic shock.

**Methods:** We conducted a multi-center retrospective cohort study of adult ICU patients with septic shock treated throughout the UPMC Health System from January 2021 to January 2022. Patients who received norepinephrine as a first-line vasopressor followed by either vasopressin or an alternative vasopressor were included. Generalized linear models with a log link and gamma distribution were used to compare adjusted total healthcare costs. Recycled predictions were used to estimate marginal cost differences. GOSSIS performance was evaluated by comparing predicted versus observed ICU mortality across risk groups.

**Results:** Among 3,898 eligible patients, 1,060 received vasopressin and 2,838 received alternative second-line vasopressors. Vasopressin use was associated with significantly higher adjusted hospital costs (cost ratio = 1.14, 95% CI: 1.08, 1.20). The estimated marginal cost difference was $5,887 per patient. Drug costs accounted for a greater proportion of total hospitalization costs among vasopressin users (20% vs. 12%). GOSSIS demonstrated strong calibration, with predicted ICU mortality closely aligning with observed rates across all severity strata.

**Conclusions:** Vasopressin use in septic shock was associated with a 14% increase in hospitalization costs compared to alternative second-line vasopressors. The GOSSIS demonstrated strong predictive validity for ICU mortality, supporting its potential use in guiding resource allocation decisions in critically ill populations.

## Background

Septic shock remains a critical challenge in intensive care, characterized by profound systemic inflammation, hypotension, and organ dysfunction that often require aggressive management strategies.^1^ Vasoactive agents play a central role in restoring blood pressure and vascular tone in patients who do not adequately respond to initial fluid resuscitation. According to the 2021 Surviving Sepsis Campaign International Guidelines, norepinephrine is the recommended first-line vasopressor for the treatment of septic shock.^2^ When shock is refractory to norepinephrine alone, vasopressin is recommended as an adjunct therapy to increase mean arterial pressure without further escalating catecholamine doses.^3^

Vasopressin offers distinct pharmacologic advantages through selective V1 receptor activation, which increases vascular tone without significantly raising heart rate.^4^ This mechanism can be particularly beneficial in patients where minimizing cardiac stress is a priority. Moreover, vasopressin may reduce the need for high-dose catecholamines, thereby potentially decreasing adverse effects such as tachyarrhythmias and myocardial stress.^5^ One rationale for its use in septic shock is the potential to mitigate complications associated with excessive adrenergic stimulation from agents like dopamine.^6^ However, despite these potential advantages, studies and comprehensive meta-analyses, including those with individual patient data, have consistently shown no mortality benefit compared to other second-line vasopressors and no clear evidence for subgroup-specific clinical advantages.^5,7,8^ While some earlier studies suggested possible reductions in renal replacement therapy (RRT) use or fewer arrhythmias in select subgroups, these findings have not been consistently demonstrated across analyses.^8^

In addition to these equivocal clinical outcomes, vasopressin use has also been challenged by significant economic and policy concerns. Under the FDA’s Unapproved Drugs Initiative, Par Pharmaceuticals obtained formal approval for vasopressin, which had been used for decades without such approval.^9^ This regulatory maneuver, alongside an aggressive patenting strategy, allowed Par Pharmaceuticals to raise vasopressin’s price and potentially delay some formulations of the generic until 2035.^9^ In fact, from 2022 to 2023, the Department of Health and Human Services found vasopressin to have the highest drug price increase of 3,558%.^10^ These developments had profound financial implications for hospitals, with vasopressin costs in the U.S. alone reaching nearly $600 million recently.^11^ As a result, healthcare institutions initiated cost-saving measures, including adjusting drug formulations and revising storage practices, to mitigate these burdens.^12^ Although recent entry of specific generic formulations has partially alleviated these concerns, the history of price fluctuations underscores the need to continuously assess both economic and clinical outcomes when using high-cost therapies like vasopressin.

In light of these mixed clinical outcomes and fluctuating acquisition costs, tools that can better identify patients most likely to benefit from specific therapies are increasingly important. To optimize both clinical and economic decision-making in the intensive care unit (ICU), predictive tools have gained prominence. One such tool is the Global Open-Source Severity of Illness Score (GOSSIS), a recently developed and highly scalable model designed to predict ICU mortality using large-scale, high-dimensional data from diverse patient populations.^13^ Compared to traditional scoring systems such as APACHE II, SAPS II, and MPM, which rely heavily on static variables from the first 24 hours of ICU admission, GOSSIS aims to enhance prognostic accuracy and generalizability across heterogeneous settings.^14–16^ By better characterizing patient risk, GOSSIS may support resource allocation decisions, particularly for high-cost therapies such as vasopressin, where clinical benefits must be weighed carefully against economic impact.

Given the substantial financial implications of vasopressor selection in septic shock, a better understanding of the cost differences associated with vasopressin use is critical for hospital resource planning and clinical guideline development. In parallel, risk prediction tools such as the GOSSIS may help contextualize treatment decisions by identifying patients most likely to benefit from specific interventions. To address both the economic and clinical dimensions of vasopressin use, this study primarily aimed to conduct a cost analysis comparing vasopressin to other second-line vasopressors in patients with septic shock. A secondary objective was to evaluate how well the GOSSIS performed in predicting ICU mortality within a septic shock patient population.

## Methods

### Study Design and Data Source

This retrospective cohort study analyzed patient data from 25 ICUs across11 hospitals within the UPMC healthcare system. Data were collected from January 2021 to December 2022. Only patients with complete hospitalization costs per admission data were included in the study. The study was evaluated by the UPMC Quality Review Committee and determined to meet criteria of quality improvement, which did not require Institutional Review Board approval.

### Samples and Cohort

Hospitals and ICUs within the health system were included if data were readily available with the UPMC Clinical Analytics Database and complete cost data was able to be retrieved. Patients who had at least one medication charge for a vasopressor (norepinephrine, epinephrine, dopamine, phenylephrine or vasopressin during an ICU admission were identified and evaluated for any sepsis related ICD-10 diagnosis code. Patients who were less than 18 years of age, died within 24 hours of ICU admission, had an ICU length of stay less than 24 hours, had missing cost data, or had missing GOSSIS were excluded from the study. Only patients who received norepinephrine as first-line treatment, followed by vasopressin or an alternative vasopressors as second-line therapy, were included (Figure 1). This ensured consistency in initial treatment protocols, as norepinephrine is the recommended first-line vasopressor for septic shock.^2^ Throughout the entire study period, an internal guideline with subsequent clinical decision support system alert recommended that vasopressin infusions not to be initiated until norepinephrine reached a dose threshold of 0.7 mcg/kg/min. Patients were then divided into two cohorts based on secondary vasopressor treatment: vasopressin versus an alternative vasopressor, which included epinephrine, dopamine, and phenylephrine.

**Figure 1.**
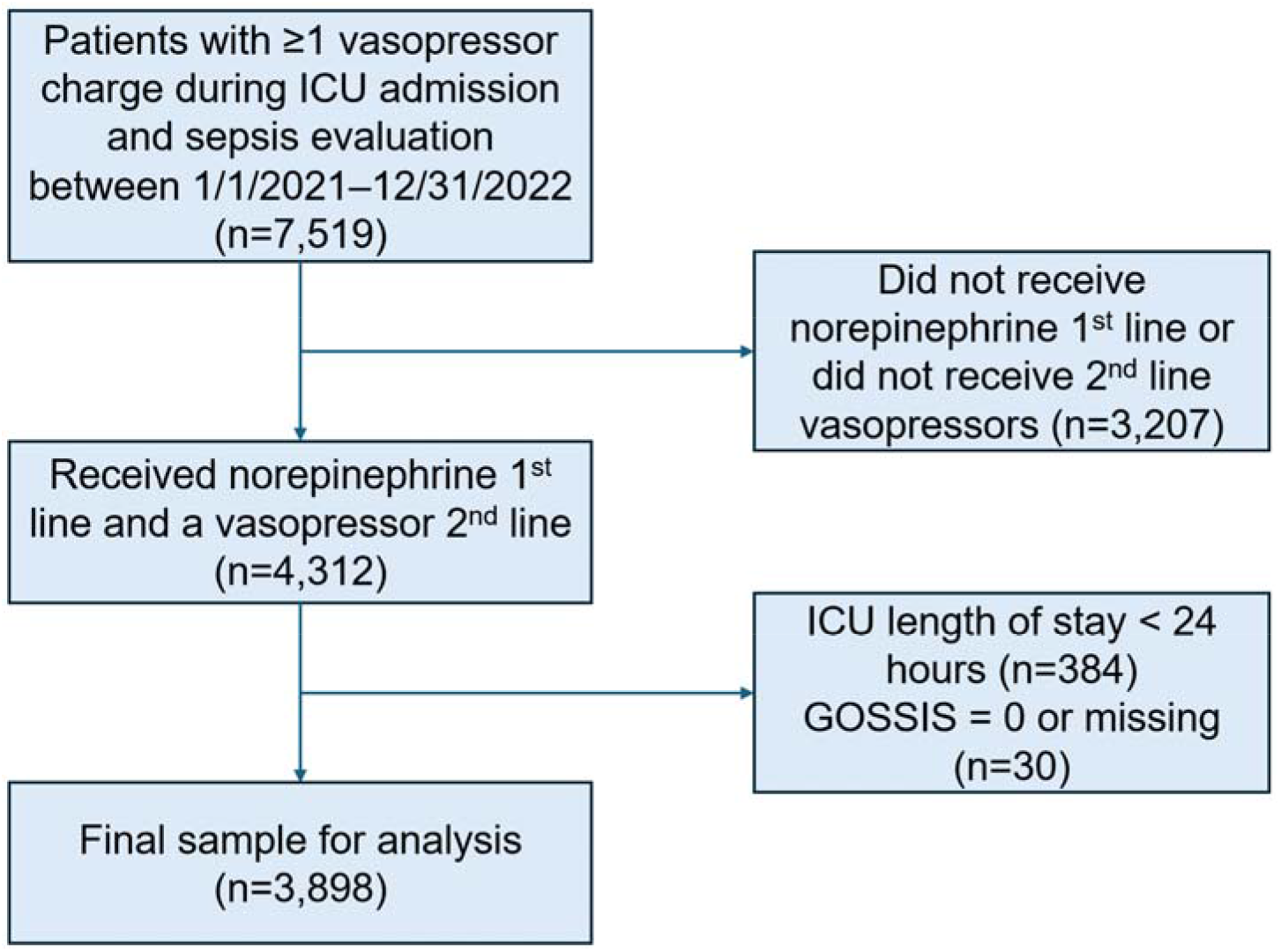
**Patient flow diagram of the study**

### Main Outcomes and Measures

The outcome of the cost analysis portion of the study was total hospitalization costs of the patient’s hospitalization stay. Total hospitalization costs were defined as the sum of direct expenses (medications, blood products, laboratory testing, etc.) and service costs (expenses related to providers throughout the hospitalization). Supporting costs such as hospital overhead were not included in the cost analysis. Costs were adjusted to 2024 U.S. dollars using the annual medical care component of the Consumer Price Index to reflect changes in healthcare inflation.^17^

For the secondary analysis of assessing the GOSSIS as a predictor of ICU mortality involved comparing the predicted mortality to the actual observed mortality for each GOSSIS group. GOSSIS was calculated upon ICU admission for all patients. Groups were created based on the GOSSIS as follows: Group 1: < 20%; Group 2: 20% to 39.9%; Group 3: 40% to 59.9%; Group 4: ≥ 60%.

### Statistical Analysis

Baseline characteristics were compared using chi-square tests for categorical data and t-tests for continuous data. Costs were analyzed using generalized linear models (GLMs) with a log link function and gamma distribution. The analysis controlled for patient demographics, hospital attributes, and clinical characteristics that could be related to healthcare costs. Recycled predictions were used to estimate the marginal (i.e. incremental costs) attributable to vasopressin usage.^18,19^ Costs were compared between patients with and without vasopressin as a cost ratio and recycled predictions produced predicted mean costs for both groups. As a sensitivity analysis, patients with the highest 1% of costs were excluded.

The secondary analysis assessed how well the GOSSIS predicted ICU mortality. Logistic regression was used to compare the odds of ICU mortality based on the GOSSIS, controlling for the previously mentioned patient and treatment characteristics. Recycled predictions were then used to calculate predicted probabilities at group levels of the GOSSIS scores, which were compared to actual mortality probabilities. All analyses were conducted using SAS 9.4 (SAS Institute Inc., Cary, NC) and Stata version 18 (Stata Corp. LP, College Station, TX).

## Results

### Baseline Characteristics

Within the eligible cohort, there were 2,838 alternative vasopressor users compared to 1,060 vasopressin users (Table 1). Among the patients treated with an alternative vasopressor, 1299 were given epinephrine (45.77%), 1204 were given phenylephrine (42.42%), and 335 were given dopamine (11.80%). Patients in both groups were close in age (65.1 years for vasopressin vs. 65.2 years for alternative vasopressor). Patients treated with vasopressin were more likely to be in quaternary care hospitals (54.8% vs. 49.2%) and less likely in community hospitals (8.3% vs. 14.1%). Additionally, a higher proportion of vasopressin-treated patients were found in medical and cardiothoracic ICUs. The distribution of GOSSIS indicated slightly higher severity scores in the vasopressin group.

**Table 1.**
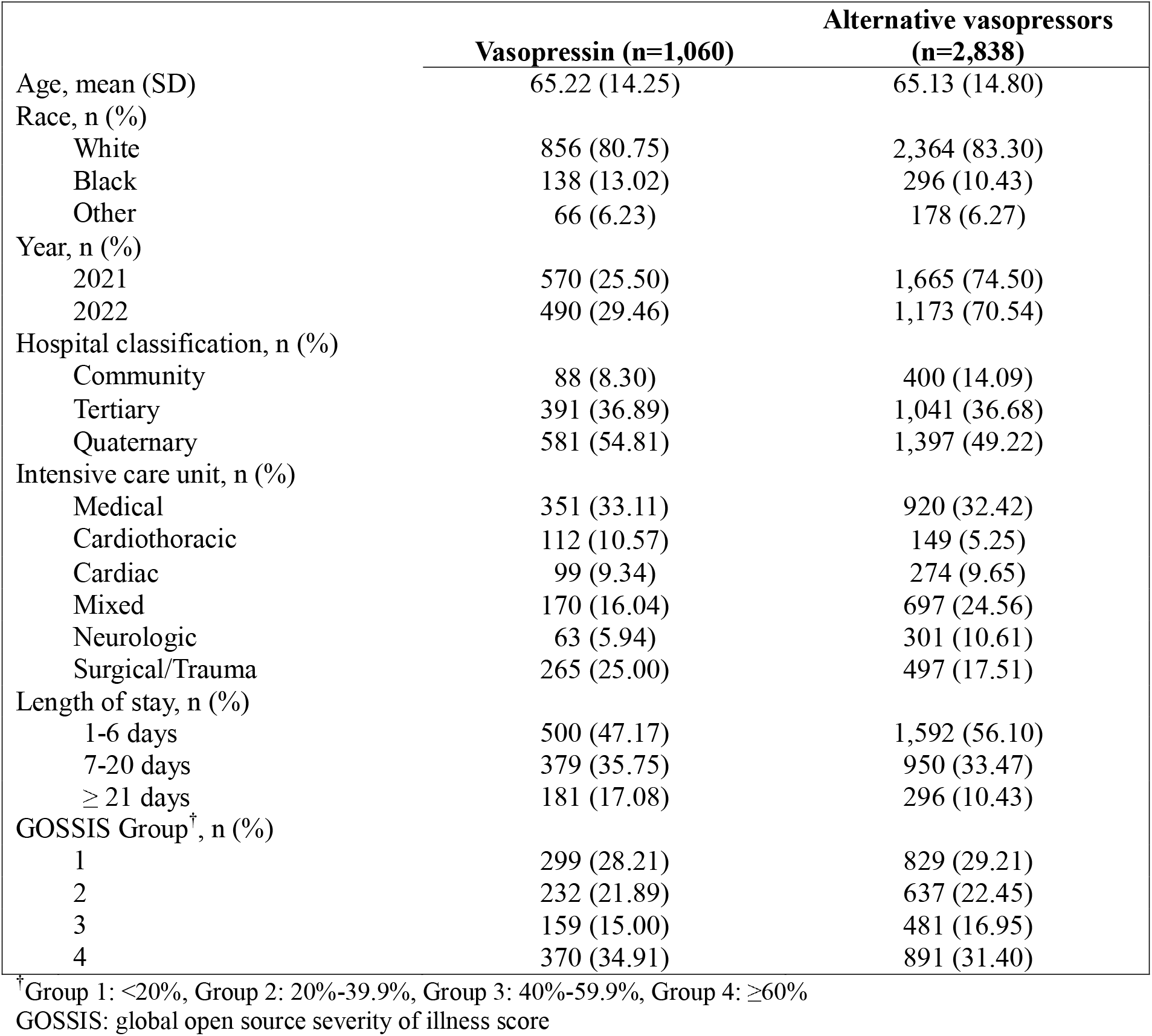
Baseline characteristics.

### Hospitalization Cost Analysis Associated with Vasopressin Use

The unadjusted mean cost for patients treated with vasopressin was $59,656 (standard deviation [SD]: 87,982) compared to $38,961 (SD: 48,986) for those treated with an alternative vasopressor. In the adjusted cost analysis, patients treated with vasopressin had 14% higher healthcare costs (cost ratio [CR]: 1.14, 95% CI: 1.08, 1.20, p<0.001) during their ICU admission compared to those treated with an alternative vasopressor (Table 2). Patient age and year of treatment did not significantly influence the cost. However, hospital setting was significant as patients treated in quaternary hospitals incurred significantly higher costs, with a cost ratio of 1.47 (95% CI: 1.35, 1.61, p<0.001) compared to those in community hospitals.

**Table 2.** Adjusted healthcare cost ratios by vasopressor use and patient characteristics.

|  | Cost ratio | 95% CI | p-value |
|---|---|---|---|
| Vasopressin | 1.14 | [1.08, 1.20] | <0.001 |
| Age (per additional decade) | 0.99 | [0.97, 1.01] | 0.59 |
| Race |  |  |  |
| White | Reference |  |  |
| Black | 0.92 | [0.86, 0.99] | <0.1 |
| Other | 1.13 | [0.99, 1.29] | 0.06 |
| Year |  |  |  |
| 2021 | Reference |  |  |
| 2022 | 0.99 | [0.90, 1.09] | 0.86 |
| Hospital classification |  |  |  |
| Community | Reference |  |  |
| Tertiary | 1.06 | [0.97, 1.16] | 0.21 |
| Quaternary | 1.47 | [1.35, 1.61] | <0.001 |
| Intensive care unit |  |  |  |
| Medical | Reference |  |  |
| Cardiothoracic | 1.31 | [1.17, 1.46] | <0.001 |
| Cardiac | 0.99 | [0.89, 1.10] | 0.90 |
| Mixed | 0.85 | [0.79, 0.91] | <0.001 |
| Neurologic | 0.93 | [0.85, 1.02] | 0.12 |
| Surgical/Trauma | 1.19 | [1.10, 1.28] | <0.001 |
| Length of stay |  |  |  |
| 1-6 days | Reference |  |  |
| 7-20 days | 2.41 | [2.29, 2.54] | <0.001 |
| ≥ 21 days | 5.85 | [5.43, 6.30] | <0.001 |
| GOSSIS Group |  |  |  |
| 1 | Reference |  |  |
| 2 | 0.96 | [0.89, 1.03] | 0.25 |
| 3 | 0.96 | [0.89, 1.05] | 0.39 |
| 4 | 0.97 | [0.90, 1.05] | 0.48 |

<sup>†</sup>Group 1: <20%, Group 2: 20%-39.9%, Group 3: 40%-59.9%, Group 4: ≥60%
GOSSIS: global open source severity of illness score

Using recycled predictions, the predicted mean costs for patients treated with vasopressin was $48,440 (95% CI: $46,085, $50,796) compared to $42,553 (95% CI: $41,039, $44,067) for those treated with an alternative vasopressor. The resultant costs attributable to using vasopressin compared to an alternative vasopressor was an additional $5,887 (95% CI: $3,402, $8,372), which was significant. After removing patients who made up the top 1% of costs as a sensitivity analysis, treatment with vasopressin was still significantly associated with higher costs (cost ratio=1.11, 95% CI: 1.06, 1.16) translating to an addition $4,250 per patient (95% CI: $2,221, $6,278).

We explored whether the higher overall costs in vasopressin-treated patients could be attributed to increased overall drug expenditures specifically. We found that drug costs accounted for 20% (SD=11.44%) of total hospital costs in vasopressin-treated patients, compared to 12% (SD=8.72%) in the alternative vasopressor group. The distribution of this proportion was right-skewed in both groups, with a longer upper tail among vasopressin users (Figure 2). Visual inspection of the smoothed density curves demonstrated greater variability in the vasopressin group indicating more heterogeneity in cost contributions.

**Figure 2.**
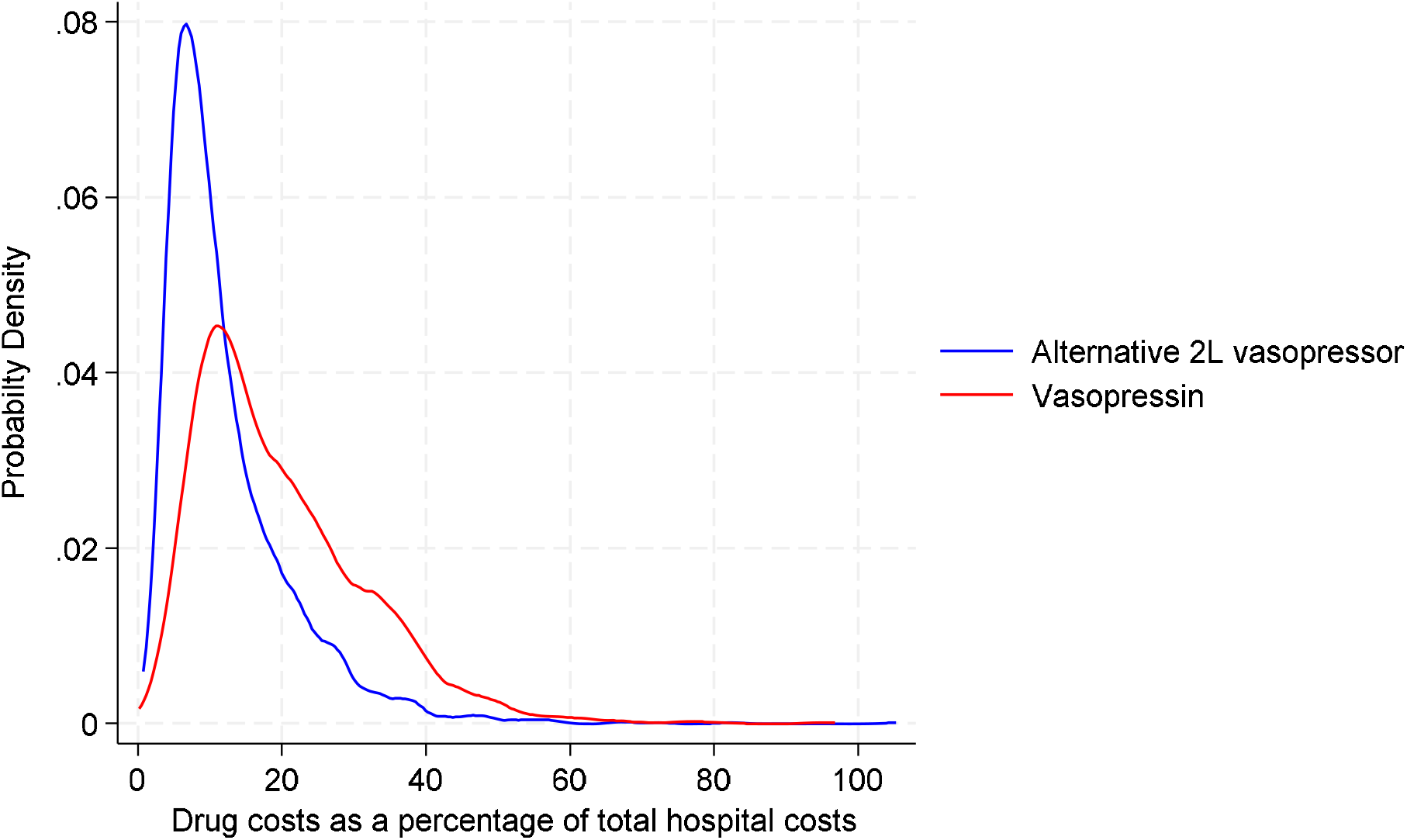
**Distribution of drug costs proportional to total hospital costs**

### GOSSIS and ICU Mortality

Our secondary analysis evaluated the GOSSIS as a predictor of ICU mortality. The results from indicated that the odds of ICU mortality increased progressively with higher GOSSIS groups when compared to group 1 (Table 3). Compared to group 1, group 2 showed a 95% increase in the odds (95% CI: 1.61, 2.35) of mortality risk, group 3 a 173% increase (95% CI: 2.22, 3.35), and group 4 a 694% increase (95% CI: 6.58, 9.58).

**Table 3.** Adjusted odds ratios for ICU mortality by GOSSIS group and vasopressor use.

|  | Odds ratio | 95% CI | p-value |
|---|---|---|---|
| Vasopressin | 1.02 | [0.87, 1.20] | 0.80 |
| Age (per additional decade) | 1.08 | [1.03, 1.14] | <0.01 |
| Race |  |  |  |
| White | Reference |  |  |
| Black | 1.04 | [0.83, 1.31] | 0.72 |
| Other | 1.65 | [1.22, 2.22] | <0.01 |
| Year |  |  |  |
| 2021 | Reference |  |  |
| 2022 | 0.91 | [0.70, 1.20] | 0.51 |
| Hospital classification |  |  |  |
| Community | Reference |  |  |
| Tertiary | 1.07 | [0.82, 1.40] | 0.60 |
| Quaternary | 0.98 | [0.75, 1.27] | 0.86 |
| Intensive care unit |  |  |  |
| Medical | Reference |  |  |
| Cardiothoracic | 1.03 | [0.77, 1.39] | 0.81 |
| Cardiac | 0.87 | [0.65, 1.16] | 0.35 |
| Mixed | 0.90 | [0.73, 1.11] | 0.33 |
| Neurologic | 0.86 | [0.66, 1.11] | 0.24 |
| Surgical/Trauma | 0.98 | [0.80, 1.19] | 0.83 |
| Length of stay |  |  |  |
| 1-6 days | Reference |  |  |
| 7-20 days | 0.97 | [0.84, 1.13] | 0.72 |
| ≥ 21 days | 1.02 | [0.82, 1.28] | 0.83 |
| GOSSIS Group |  |  |  |
| 1 | Reference |  |  |
| 2 | 1.95 | [1.61, 2.35] | <0.001 |
| 3 | 2.73 | [2.22, 3.35] | <0.001 |
| 4 | 7.94 | [6.58, 9.58] | <0.001 |

<sup>†</sup>Group 1: <20%, Group 2: 20%-39.9%, Group 3: 40%-59.9%, Group 4: ≥60%
GOSSIS: global open source severity of illness score

The predicted ICU mortality rates from the GOSSIS model closely matched the observed mortality across all four risk groups (Table 4). For example, group 2 had a predicted mortality of 45.14% (95% CI: 41.81%, 48.47%) and an observed mortality of 45.11%, while group 3 showed a predicted mortality of 53.59% (95% CI: 49.68%, 57.50%) and an observed rate of 54.22%. In group 1, the predicted mortality was 29.72%, compared to 28.90% observed. In group 4, predicted and observed mortality were 77.05% and 77.40%, respectively. The predicted and observed values were similar in all groups, with observed rates falling within the 95% confidence intervals of the model-based estimates.

**Table 4.** Predicted vs. observed ICU mortality by GOSSIS group.

| GOSSIS Group | **Predicted mortality (95% CI)** | **Observed mortality** |
|---|---|---|
| 1 | 29.72% (26.99%, 32.46%) | 28.90% (n=326/1,128) |
| 2 | 45.14% (41.81%, 48.47%) | 45.11% (n=392/869) |
| 3 | 53.59% (49.68%, 57.50%) | 54.22% (n=347/640) |
| 4 | 77.05% (74.70%, 79.39%) | 77.40% (n=976/1,261) |

<sup>†</sup>Group 1: <20%, Group 2: 20%-39.9%, Group 3: 40%-59.9%, Group 4: ≥60%
GOSSIS: global open source severity of illness score

## Discussion

This study provides the first comprehensive pragmatic cost analysis comparing vasopressin with alternative second-line vasopressors in the treatment of septic shock. We found that vasopressin use was associated with a 14% increase in controllable hospitalization costs, a finding consistent with the well-documented surge in its acquisition price following market rebranding. Although our adjusted models accounted for patient characteristics, ICU type, and hospital-level factors, cost differences persisted. To better understand whether this difference was primarily driven by vasopressin’s acquisition price versus broader markers of care intensity, we examined drug costs as a proportion of total hospitalization costs. Vasopressin-treated patients had a higher share of costs allocated to drug expenditures (20% vs. 12%) suggesting that drug-specific spending itself may be a meaningful contributor to the higher overall costs associated with vasopressin use. This reinforces the need to weigh acquisition cost and stewardship strategies when considering vasopressin in ICU practice.

Although at the time of this study vasopressin was associated with higher acquisition costs, these figures should be interpreted within the broader clinical and economic context. Prior studies have suggested that vasopressin may reduce downstream healthcare utilization, including shorter ICU stays and reduced need for renal replacement therapy, which could partially offset initial drug costs.^20^ Additionally, demand for vasopressin has appeared relatively inelastic—usage continued to rise even after an average per-patient price increase of $527 following rebranding, reflecting the clinical belief that vasopressin’s physiologic advantages justify its cost.^21^ Institutional-level initiatives, such as modifying formulation concentrations, removing vasopressin from automated dispensing cabinets, or optimizing storage, have shown potential to lower drug waste and reduce costs without compromising care.^12,22^ For example, a previous study reported approximately $235,000 in annual savings following such interventions at a large academic center.^12^ These findings underscore the importance of site-specific stewardship strategies, especially for high-cost therapies like vasopressin.

Our analysis also identified substantial variation in vasopressor-associated hospitalization costs across care settings. Patients treated in quaternary care hospitals experienced significantly higher costs, consistent with prior work linking academic and high-acuity centers to elevated ICU expenditures.^23^ Higher costs in these settings may reflect increased use of advanced monitoring, mechanical support, and more resource-intensive protocols. Similarly, patients managed in cardiothoracic and surgical/trauma ICUs had higher cost ratios, which aligns with prior literature identifying these ICU types as high-cost environments due to frequent use of organ support modalities.^24^ Although our dataset did not capture device-level interventions, such measures are among the strongest predictors of ICU spending.^23^ These variations emphasize the role of institutional context, specialty practice patterns, and care intensity in shaping cost trajectories. Importantly, ICU-level resource use has been shown to vary substantially across institutions without corresponding improvements in adjusted mortality, suggesting opportunities for optimizing both clinical outcomes and economic efficiency.^25^

In terms of clinical effectiveness, vasopressin use was not associated with a significant difference in ICU mortality compared to alternative second-line vasopressors (OR=1.01, 95% CI: 0.86, 1.18). This aligns with meta-analyses and randomized trials showing that while vasopressin may reduce the incidence of arrhythmias or need for renal replacement therapy, it does not confer a consistent survival advantage.^7,8^ Some studies have proposed that earlier initiation of vasopressin may improve hemodynamic outcomes or reduce the duration of organ support, though whether these translate into long-term mortality or cost benefits remains uncertain.^26–28^ Given these mixed findings, decisions regarding vasopressor choice should integrate both physiologic considerations and broader system-level implications, including cost, stewardship infrastructure, and formulary flexibility.

Beyond the cost and mortality findings, our study also confirmed the strong performance of the GOSSIS model in our ICU population. Predicted ICU mortality closely matched observed outcomes across all four severity strata, with all observed values falling within the 95% confidence intervals of model-based estimates. This high level of calibration supports the validity of GOSSIS as a risk stratification tool across diverse ICU settings. While SOFA remains widely used for organ dysfunction scoring, GOSSIS incorporates a broader set of physiologic and laboratory variables and may offer more granular risk estimation in some contexts.^13,29^ When combined with cost data, risk stratification tools like GOSSIS may help guide ICU resource allocation and inform quality benchmarking efforts. For example, integrating predictive models with cost data may enable hospitals to identify patients at high risk for poor outcomes and high-cost trajectories, ultimately supporting value-based care initiatives.

Taken together, our findings highlight the importance of aligning clinical decision-making with economic realities in the ICU. Vasopressin remains a pharmacologically appealing agent, but its historical pricing and variation in usage across settings underscore the need for ongoing cost-effectiveness evaluations. As generic formulations gain market share and policy efforts to reform the Unapproved Drugs Initiative progress, the economic landscape surrounding vasopressin will likely evolve. Future research should continue to evaluate the clinical and economic trade-offs of vasopressor strategies, incorporating both patient-centered outcomes and system-level cost data to guide stewardship and formulary decisions in critical care.

This study had several limitations. Despite adjustment for multiple clinical and demographic factors, residual confounding is possible, especially regarding unmeasured variables such as vasopressor selection rationale or timing. The analysis was limited to a single healthcare system, which may reduce generalizability, particularly given system-specific guidelines for use of vasopressors. Cost estimates excluded long-term healthcare expenses, and acquisition prices reflect historical rather than current costs. Additionally, specific vasopressor drug costs could not be isolated, though we addressed this in part by comparing drug cost proportions across groups. Finally, we were unable to evaluate vasopressin-related outcomes such as arrhythmias or renal replacement therapy. Future studies should assess long-term costs and outcomes associated with protocolized vasopressin use.^30–32^

## Conclusion

In conclusion, vasopressin use as a second-line agent in septic shock was associated with higher hospitalization costs. While vasopressin may offer pharmacologic advantages in select clinical contexts, its cost implications warrant careful consideration. The GOSSIS model demonstrated strong calibration and proved useful for risk stratification in critically ill patients. These results contribute to the growing evidence base guiding vasopressor selection and emphasize the importance of integrating clinical and economic data into critical care decision-making.

## Data Availability

All data produced in the present work are contained in the manuscript

## Notes

### Competing Interest Statement

The authors have declared no competing interest.

